# Rapid waning of protection induced by prior BA.1/BA.2 infection against BA.5 infection

**DOI:** 10.1101/2022.08.16.22278820

**Authors:** João Malato, Ruy M. Ribeiro, Eugénia Fernandes, Pedro Pinto Leite, Pedro Casaca, Carlos Antunes, Válter R. Fonseca, Manuel Carmo Gomes, Luis Graca

**Author notes:** **Corresponding author:** Luis Graca MD DPhil. Instituto de Medicina Molecular João Lobo Antunes, Faculdade de Medicina, Centro Académico de Medicina de Lisboa, Universidade de Lisboa. Avenida Professor Egas Moniz, 1649-028 Lisboa, Portugal. Joint senior authors (Manuel Carmo Gomes, Luis Graca).

## Abstract

SARS-CoV-2 omicron subvariants BA.1 and BA.2 became dominant in many countries in early 2022. These subvariants are now being displaced by BA.4 and BA.5. While natural infection with BA.1/BA.2 provides some protection against BA.4/BA.5 infection, the duration of this protection remains unknown.

We used the national Portuguese COVID-19 registry to investigate the waning of protective immunity conferred by prior BA.1/BA.2 infection towards BA.5. We divided the individuals infected during the period of BA.1/BA.2 dominance (>90% of sample isolates) in successive 15-day intervals and determined the risk of subsequent infection with BA.5 over a fixed period.

Compared with uninfected people, one previous infection conferred substantial protection against BA.5 re-infection at 3 months (RR=0.12; 95% CI: 0.11-0.12). However, although still significant, the protection was reduced by two-fold at 5 months post-infection (RR=0.24; 0.23-0.24).

These results should be interpreted in the context of vaccine breakthrough infections, as the vaccination coverage in the individuals included in the analyses is >98% since the end of 2021.

This waning of protection following BA.1/BA.2 infection highlights the need to assess the stability and durability of immune protection induced with the adapted vaccines (based on BA.1) over time.

---

The SARS-CoV-2 omicron BA.5 subvariant has been progressively displacing earlier BA.1 and BA.2 subvariants. We and others have recently reported that natural infection with BA.1/BA.2 offers significant protection against BA.5 reinfection^1–3^. However, the duration of the protective effect is a concern, given the poor ability of BA.1/BA.2 infection to achieve robust neutralizing antibodies against BA.5^4,5^. This issue is critical as the current adapted omicron vaccines under development are based on BA.1 ^6^.

Portugal was one of the first countries affected by omicron BA.1 and, more recently, omicron BA.5. We used national SARS-CoV-2 genetic surveillance data to identify periods when different variants represented >90% of the isolates^1,7^. Then, we used the Portuguese COVID-19 registry (*SINAVE*), which includes all notified cases of infection in the country based on a positive molecular or professional rapid antigen test for SARS-CoV-2 and irrespective of clinical presentation, to identify all individuals infected during those periods. Reinfections were defined as two positive tests in the same individual with an interval of at least 90 days^8^. Furthermore, our study included all reported cases in individuals 12 years and older in Portugal, for whom vaccination coverage was >98% by the end of 2021. Thus, the results ought to be interpreted as related to breakthrough infections in a highly vaccinated population.

To investigate whether there is evidence for waning protection of a previous BA.1/BA.2 infection against BA.5, we divided the period of BA.1/BA.2 dominance into four time periods (each with 15 days), and the time with pre-omicron variants (2020 and 2021) into 3-month intervals (Figure 1A). We found that protection against BA.5, among those previously infected with BA.1/BA.2 (and no other recorded infection), was higher for individuals with more recent infections (Figure 1A, Table 1). Indeed, protection was reduced over a 2-month period from 88% (based on 1-RR) at 3 months to 76% at 5 months. Furthermore, protection against BA.5 acquired from a single pre-omicron infection was consistently lower and stabilized at about 50% (Figure 1A).

**Figure 1.**
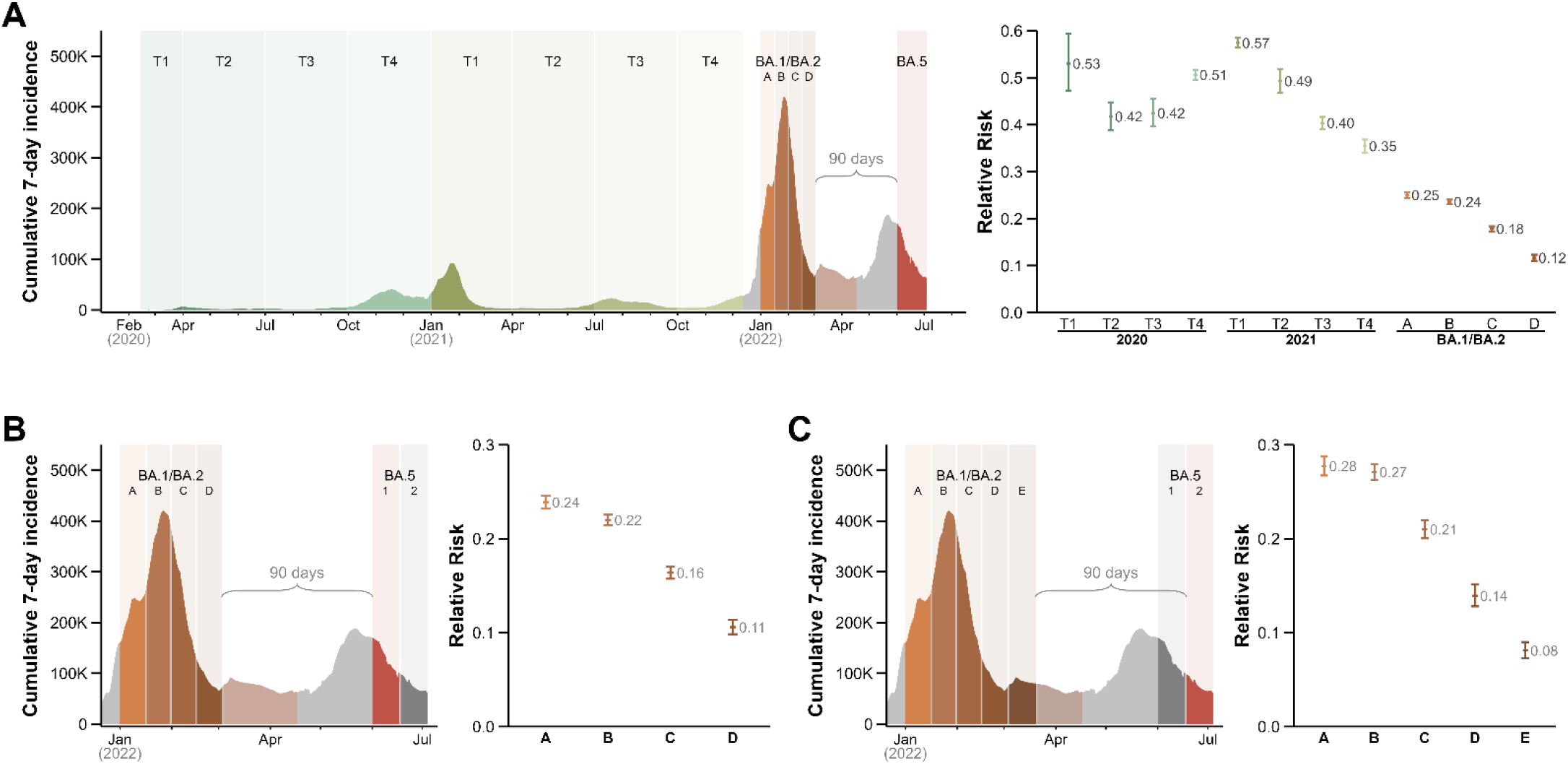
Waning of the protective effect of a single previous infection on omicron BA.5. **A**. We identified periods (in different colors) where one variant was represented in >90% of sample isolates (data from the national SARS-CoV-2 genetic diversity surveillance^4^). During the periods in grey no single variant dominated infection. We divided the period of BA.1/BA.2 dominance into four 15-day intervals. We calculated the relative risk (RR) of infection during the period of BA.5 dominance (June 1^st^ to July 4^th^, in red) for individuals with one previous infection in the periods of dominance of different variants, and in the four intervals of BA.1/BA.2 dominance (A to D), but without any further documented infection until June 1^st^. RR of infection for individuals with one previous infection at different periods during 2020 and 2021, and in the four 15-day periods of BA.1/BA.2 dominance (right). **B**. RR of infection during the 1^st^ period of BA.5 dominance (June 1^st^ – 17^th^) for individuals infected in the four intervals of BA.1/BA.2 dominance. **C**. RR of infection during the 2^nd^ period of BA.5 dominance (June 18^th^ – July 4^th^) for individuals infected in the five represented intervals of BA.1/BA.2 dominance.

**Table 1.** Risk of omicron BA.5 infection for individuals infected with BA.1/BA.2 at different intervals. We included in the study the population 12 years and older. Under “1^st^ infection” is the number of individuals at risk for a second infection by BA.5 (i.e., without a second infection before June 1^st^ or June 17^th^). Reinfections were defined as two positive tests by the same individual more than 90 days apart. Note that the risk is dependent on the epidemic situation and differs in the two BA.5 periods. Tri, trimester; inf, infection; RR, relative risk; CI, confidence interval.

|  | Date interval | Uninfected on<br>June 1<sup>st</sup> 2022 | 1<sup>st</sup><br>infection | BA.5<br>infection | Absolute<br>Risk | RR (95% CI) |
| --- | --- | --- | --- | --- | --- | --- |
| Uninfected | – | 5 328 287 | – | 367 783 | 0.069 | – |
| 2020 (Tri. 1) | 14/02/20 – 31/03/20 | – | 8 101 | 306 | 0.038 | 0.530 (0.473, 0.594) |
| (Tri. 2) | 01/04/20 – 30/06/20 | – | 26 379 | 789 | 0.030 | 0.417 (0.389, 0.448) |
| (Tri. 3) | 01/07/20 – 30/09/20 | – | 26 644 | 811 | 0.030 | 0.425 (0.396, 0.455) |
| (Tri. 4) | 01/10/20 – 31/12/20 | – | 253 453 | 8 958 | 0.035 | 0.506 (0.496, 0.517) |
| 2021 (Tri. 1) | 01/01/21 – 31/03/21 | – | 292 731 | 11 683 | 0.040 | 0.574 (0.564, 0.585) |
| (Tri. 2) | 01/04/21 – 30/06/21 | – | 44 064 | 1 547 | 0.035 | 0.493 (0.469, 0.518) |
| (Tri. 3) | 01/07/21 – 30/09/21 | – | 133 061 | 3 807 | 0.029 | 0.403 (0.391, 0.417) |
| (Tri. 4) | 01/10/21 – 12/12/21 | – | 99 770 | 2 522 | 0.025 | 0.354 (0.341, 0.368) |
| BA.1/BA.2 (A) | 01/01/22 – 16/01/22 | – | 428 775 | 7 341 | 0.017 | 0.250 (0.244, 0.256) |
| BA.1/BA.2 (B) | 17/01/22 – 31/01/22 | – | 631 142 | 9 911 | 0.016 | 0.236 (0.231, 0.240) |
| BA.1/BA.2 (C) | 01/02/22 – 15/02/22 | – | 347 091 | 4 286 | 0.012 | 0.178 (0.173, 0.184) |
| BA.1/BA.2 (D) | 16/02/22 – 03/03/22 | – | 150 627 | 1 255 | 0.008 | 0.116 (0.110, 0.123) |
|  |  | Uninfected on<br>June 1<sup>st</sup> |  | BA.5 inf<br>1 – 17 Jun |  |  |
| Uninfected | – | 5 328 287 | – | 236 713 | 0.044 | – |
| BA.1/BA.2 (A) | 01/01/22 – 16/01/22 | – | 428 775 | 4 432 | 0.010 | 0.239 (0.232, 0.246) |
| BA.1/BA.2 (B) | 17/01/22 – 31/01/22 | – | 631 142 | 5 835 | 0.009 | 0.220 (0.214, 0.226) |
| BA.1/BA.2 (C) | 01/02/22 – 15/02/22 | – | 347 091 | 2 483 | 0.007 | 0.164 (0.157, 0.170) |
| BA.1/BA.2 (D) | 16/02/22 – 03/03/22 | – | 150 627 | 719 | 0.005 | 0.106 (0.098, 0.114) |
|  |  | Uninfected on<br>June 17<sup>th</sup> |  | BA.5 inf<br>18Jun - 4Jul |  |  |
| Uninfected | – | 5 091 574 | – | 131 070 | 0.026 | – |
| BA.1/BA.2 (A) | 01/01/22 – 16/01/22 | – | 424 343 | 2 909 | 0.007 | 0.277 (0.267, 0.287) |
| BA.1/BA.2 (B) | 17/01/22 – 31/01/22 | – | 625 307 | 4 076 | 0.007 | 0.271 (0.263, 0.279) |
| BA.1/BA.2 (C) | 01/02/22 – 15/02/22 | – | 344 608 | 1 803 | 0.005 | 0.210 (0.200, 0.220) |
| BA.1/BA.2 (D) | 16/02/22 – 03/03/22 | – | 149 908 | 536 | 0.004 | 0.139 (0.128, 0.152) |
| BA.1/BA.2 (E) | 04/03/22 – 20/03/22 | – | 165 977 | 344 | 0.002 | 0.081 (0.073, 0.090) |

To confirm that the observed waning was reproducible, we divided the period of BA.5 infection into two equal intervals (each with 17 days) and repeated the analysis for each of these two periods. The rate of protection waning was similar for the two periods of BA.5 infection with a clear ∼2 weeks delay (Figure 1B,C and Table 1). Over two months (from 3 to 5 months after the first infection), the relative risk (RR) doubled (from ∼0.12 to ∼0.25). The results also suggest that the rate of decline is faster in the initial months, with a greater change of the RR between months 3 and 4 (Figure 1B,C).

Next, we investigated whether infection with a pre-omicron variant would display a similar kinetic of waning of protection on the same timeframe. We divided the period of dominance of the Delta variant into 15-day intervals, and assessed whether individuals infected during one of those periods (and no other recorded infection) differ in their protection against infection during a period of BA.1/BA.2 dominance between Jan 1^st^ and Feb 4^th^, 2021, chosen to have equivalent duration to the BA.5 period studied above (Figure 2 and Table 2). We found similar evidence of rapid waning of protection, especially between months 3 and 4, from 80% to about 60%, respectively. Furthermore, the RR for infection with BA.1/BA.2 in individuals previously infected with the Delta variant was consistently higher (at all equivalent time intervals) when compared with the RR of omicron BA.5 infection among BA.1/BA.2-infected individuals.

**Figure 2.**
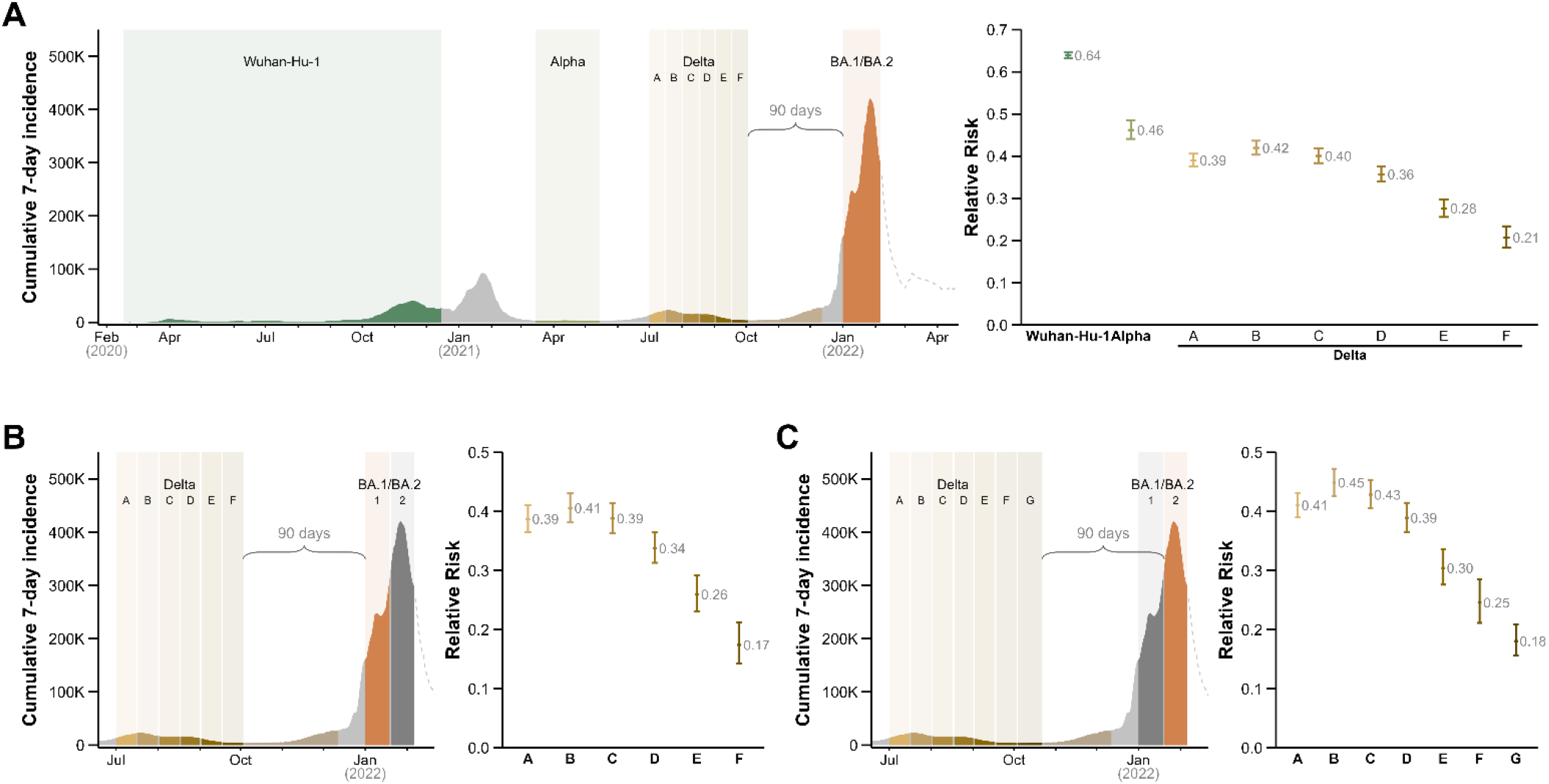
Waning of the protective effect of a single previous infection on omicron BA.1/BA.2. **A**. We identified periods (in different colors) where one variant was represented in >90% of sample isolates as described. We focused on Delta’s dominance period, up until the 90 days preceding BA.1/BA.2’s dominance, and divided it into six 15-day intervals. We calculated the relative risk (RR) of infection during the initial period of BA.1/BA.2 dominance (January 1^st^ to February 4^th^ 2022) for individuals with one infection in the periods of dominance of different variants, and in the six intervals of Delta dominance (A to F). **B** RR of infection during the 1^st^ timeslot of the BA.1/BA.2 dominance for individuals infected in the six intervals of Delta dominance. **C**. RR of infection during the 2^nd^ timeslot of the BA.1/BA.2 dominance for individuals infected in the seven represented intervals of Delta dominance.

**Table 2.** Risk of omicron BA.1 infection for individuals infected with Delta at different intervals, or previous variants. Under “1^st^ infection” is the number of individuals at risk for a second infection by BA.1/BA.2 (i.e., without a second infection before January 1^st^ 2022). Reinfections were defined as two positive tests for the same individual more than 90 days apart. RR, relative risk; CI, confidence interval.

|  | Date interval | Uninfected on<br>Jan 1<sup>st</sup> 2022 | 1<sup>st</sup><br>infection | BA.1/BA.2<br>infection | Absolute<br>Risk | RR (95% CI) |
| --- | --- | --- | --- | --- | --- | --- |
| Uninfected | – | 8 002 950 | – | 1 256 516 | 0.157 | – |
| Wuhan-Hu-1 | 17/02/20 – 15/12/20 | – | 333 780 | 34 990 | 0.105 | 0.639 (0.632, 0.646) |
| Alpha | 15/03/21 – 15/05/21 | – | 23 984 | 1 898 | 0.079 | 0.462 (0.441, 0.484) |
| Delta (A) | 01/07/21 – 16/07/21 | – | 40 238 | 2 718 | 0.068 | 0.390 (0.375, 0.406) |
| Delta (B) | 17/07/21 – 01/08/21 | – | 36 622 | 2 648 | 0.072 | 0.420 (0.403, 0.437) |
| Delta (C) | 02/08/21 – 17/08/21 | – | 32 218 | 2 232 | 0.069 | 0.401 (0.384, 0.418) |
| Delta (D) | 18/08/21 – 01/09/21 | – | 27 045 | 1 683 | 0.062 | 0.357 (0.340, 0.375) |
| Delta (E) | 02/09/21 – 17/09/21 | – | 14 220 | 695 | 0.049 | 0.276 (0.256, 0.298) |
| Delta (F) | 18/09/21 – 03/10/21 | – | 7 317 | 272 | 0.037 | 0.207 (0.184, 0.234) |

The comparison between the protection induced with BA.1/BA.2-infection towards BA.5 (Figure 1) with the protection acquired with Delta-infection against BA.1/BA.2 (Figure 2) over the same timeframe (3 – 5 months following infection) is consistent with prior reports showing infection with omicron subvariants are superior to pre-omicron variants in inducing protection against infection with other omicron subvariants^1,3,9–11^. In any case, the same trend of rapid waning especially between 3 – 4 months after infection, followed by smaller decreases in the ensuing months 4 – 5, can be observed in both cases (Delta to BA.1/BA.2; and BA.1/BA.2 to BA.5). Given the reported greater protection efficacy induced by omicron subvariants against other omicron subvariants, it is not surprising that the RR for BA.5 infection following BA.1/BA.2 exposure appears to plateau at a lower value (∼0.25) when compared with Delta to BA.1/BA.2 (∼0.40). This could be due to larger genetic difference between Delta and BA.1/BA.2 in comparison to BA.1/BA.2 and BA.5; or it could be due to different vaccine coverage during the primary infection in the studied periods of Delta or BA.1/BA.2 dominance.

A possible confounder in our study is that, although >98% of the studied population had the primary vaccine series during the BA.5 period of dominance, there was some heterogeneity in coverage with a booster dose, which was only 82%. To investigate the putative impact of a booster dose in the results, we took advantage of the fact that, in Portugal, boosters were not recommended for adolescents between 12 and 18. We thus compared the RR of reinfection for this population without boosters with the RR for the population 60 and older, which had a booster coverage >95% at the start of the BA.5 period.

We found that immune waning follows similar kinetics in the two groups of BA.1/BA.2-infected individuals, irrespective of a booster (Figure 3, Table 3), with protection being reduced from about 90% at 3 months up to 80% at 5 months. Although the absence of a clear effect of a booster dose in slowing the rate of protection waning may appear counterintuitive, given the clear benefit of booster doses in preventing severe disease, it is a result surprisingly aligned with prior reports on vaccination^12^. Indeed, it was shown that the rate of waning of vaccine effectiveness against omicron infection of a second booster (i.e., administered to individuals with a previous booster) is as fast as the waning of the first booster dose (i.e., administered to individuals that did not receive a previous booster)^12,13^. It should be stressed that our experimental design did not quantify the potential added benefit of a booster dose for protection: the design assesses rates of waning of protection following a prior infection, being the RR calculated in relation to an equivalent uninfected population (i.e., with a booster in the case of the elderly group). Of note, although a booster does not appear to reduce the waning of protection acquired following infection (according to our data) or subsequent vaccination^12,13^, booster doses were shown to significantly improve protection against severe COVID-19 following omicron BA.5 infection^14^.

**Figure 3.**
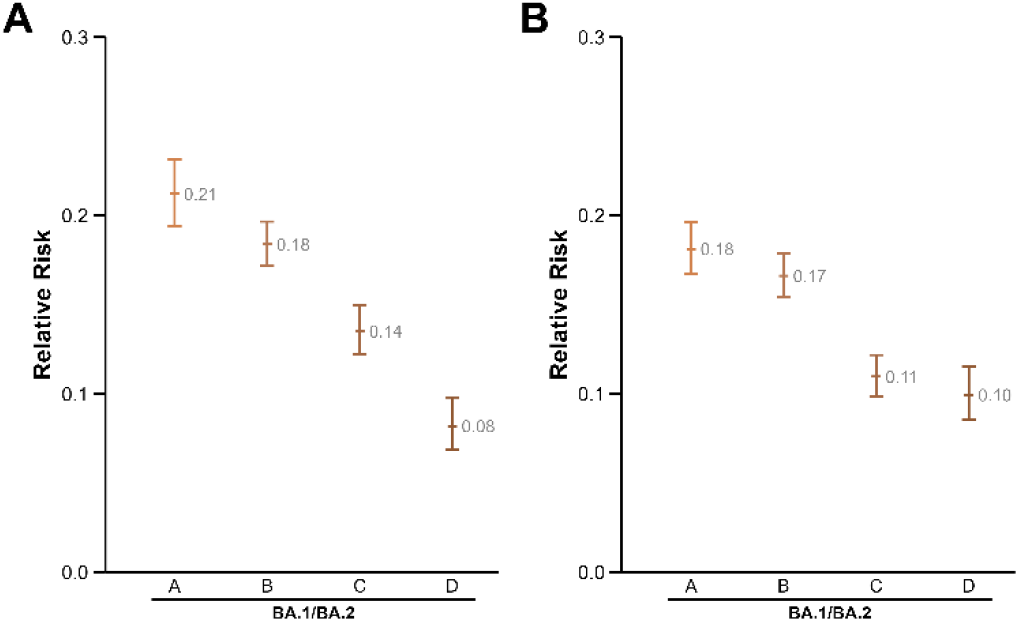
Change in the RR of BA.5 infection following exposure to BA.1/BA.2 in populations with and without a booster dose. To investigate whether vaccination with a booster dose (based on age) had an impact on the waning of protection acquired following BA.1/BA.2 infection we calculated RR of infection in two different populations. **A**. We studied the population aged 12 to 17, who were not vaccinated with a booster dose. We divided the period of BA.1/BA.2 dominance into four 15-day intervals, as described in Figure 1, and calculated the relative risk of infection during the period of BA.5 dominance (June 1st to July 4th) for individuals with one prior BA.1/BA.2 infection. **B**. We followed the same strategy for the population aged 60 and above, who had >95% coverage with a booster dose by the start of the period of BA.5 dominance. The follow-up of the two populations (12-18 and 60+) occurred simultaneously, under the same epidemiologic conditions, between June 1^st^ and July 4^th^.

**Figure 4.**
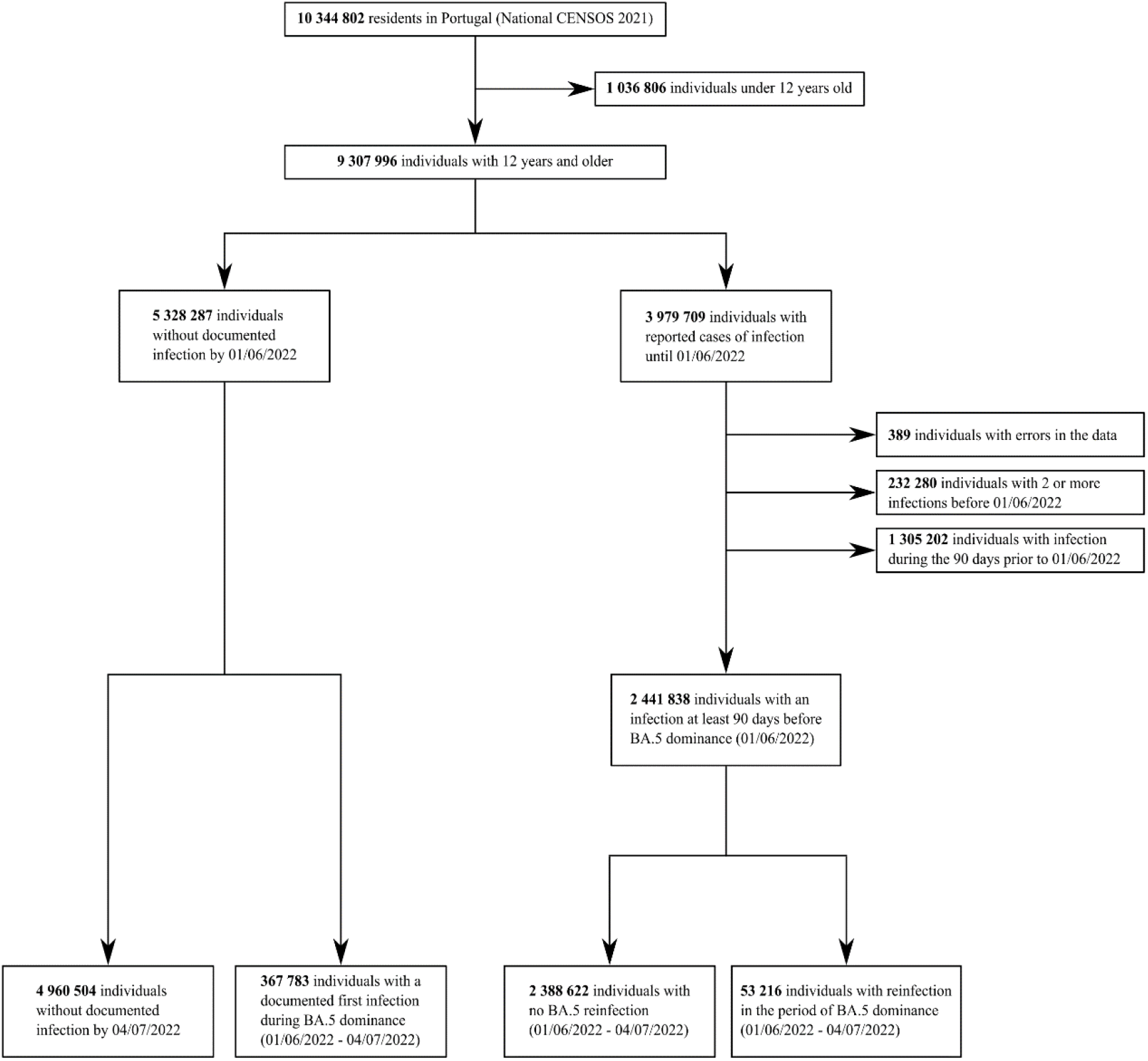
Flowchart describing the population selection. Representative flowchart, related to the population selection for the analysis presented in Figure 1A and Table 1.

**Table 3.** Risk of omicron BA.5 infection among individuals with and without a booster dose and prior infection with BA.1/BA.2 at different intervals. We compared the population aged 12 to 17, that did not receive booster doses, with individuals aged 60 years with a coverage of booster doses over 95% by June 1^st^ 2022. inf, infection; RR, relative risk; CI, confidence interval.

| 12 – 17<br>years old | Date interval | Uninfected on<br>Jun 1<sup>st</sup> 2022 | 1<sup>st</sup><br>infection | BA.5 inf | Absolute<br>Risk | RR (95% CI) |
| --- | --- | --- | --- | --- | --- | --- |
| Uninfected | – | 293 166 | – | 13 513 | 0.046 | – |
| BA.1/BA.2 (A) | 01/01/22 – 16/01/22 | – | 41 125 | 486 | 0.012 | 0.212 (0.194, 0.232) |
| BA.1/BA.2 (B) | 17/01/22 – 31/01/22 | – | 89 271 | 811 | 0.009 | 0.184 (0.172, 0.197) |
| BA.1/BA.2 (C) | 01/02/22 – 15/02/22 | – | 51 229 | 373 | 0.007 | 0.135 (0.122, 0.149) |
| BA.1/BA.2 (D) | 16/02/22 – 03/03/22 | – | 25 320 | 120 | 0.005 | 0.082 (0.069, 0.098) |
| 60 years<br>and older |  |  |  |  |  |  |
| Uninfected | – | 2 305 139 | – | 125 949 | 0.055 | – |
| BA.1/BA.2 (A) | 01/01/22 – 16/01/22 | – | 60 668 | 615 | 0.010 | 0.181 (0.167, 0.196) |
| BA.1/BA.2 (B) | 17/01/22 – 31/01/22 | – | 76 371 | 706 | 0.009 | 0.166 (0.154, 0.179) |
| BA.1/BA.2 (C) | 01/02/22 – 15/02/22 | – | 56 853 | 350 | 0.006 | 0.110 (0.099, 0.122) |
| BA.1/BA.2 (D) | 16/02/22 – 03/03/22 | – | 30 140 | 170 | 0.006 | 0.099 (0.085, 0.116) |

Our data also show that age does not significantly affect the rate of waning, as it can be inferred from the comparison between the groups aged 12-17 and 60+ (Figure 3). The similar rate of waning across different ages is not surprising, as it was previously reported that the rate of decline of neutralizing antibodies follows a similar slope irrespective of the initial titre^15^. It is likely, however, that individuals with lower initial titers will reach a threshold of poor protection sooner (given the similar slope of decline).

Registry-based studies are well suited to address immune waning. While a test-negative design may allow a more controlled comparison of a group of positive cases with negative tested controls^10^, it is in essence a retrospective case-control study. On the other hand, a registry study, such as this one, has the advantage of gathering large groups of hundreds of thousands of participants. The key issue is then to split the population into subgroups representing distinct strata and following them over the same prospective period. In fact, since the study interval (period of BA.5 dominance) is identical for all subgroups, it is even possible to calculate and compare absolute risks, which is not possible with a test-negative design.

The same reasoning can be made for other demographic characteristics that are usually hard to control for: individuals with high-risk occupations, lifestyles, or living environments may tend to be infected earlier in the pandemic. As our design allowed to test overlapping cohorts at different intervals (adjacent timeslots tested for risk in the same BA.5 period, and the RR of the same timeslot calculated in adjacent BA.5 periods), even these hard-to-control variables are unlikely to introduce significant bias in our conclusions.

One study limitation is the lack of covariate information, including symptomatic and asymptomatic infection or disease severity, as this information is absent from the records. While the groups of individuals who we followed are sufficiently large to have a relatively homogeneous distribution of symptomatic and asymptomatic cases at different time points (especially within the period of dominance of each variant), it is possible that the risk of reinfection may differ depending on the severity of the initial infection^16^. Furthermore, we are also pooling individuals with BA.5 infection, regardless of their clinical information. It is likely that protection against asymptomatic BA.5 infection wanes faster than protection against infection with clinical manifestations, as it has been repeatedly observed, for example, following vaccination^15,17^. As a consequence, the immune waning may also be different depending on the disease severity.

Overall, these data show that hybrid immunity induced following BA.1/BA.2 infection in vaccinated individuals leads to protection against BA.5 infection that wanes in the initial 5 months following infection, although retaining substantial protection efficacy at the end of those 5 months. At a time when the adapted vaccines under clinical development are based on omicron BA.1, these data show it is important to consider the rate of immune waning especially to address the potential benefit of the adapted vaccines on viral transmission.

## Methods

### Participant selection

The population included in the study comprises: (1) All individuals resident in Portugal aged 12 years and older without a documented infection until the start of the study period; (2) All individuals resident in Portugal aged 12 years and older with a single documented infection up to 90 days before the study period (see flowchart in the supplementary file, Figure 1).

We used the national COVID-19 registry (*SINAVE*) to obtain information on all notified cases of infection, irrespective of clinical presentation. The “uninfected” population was defined as the population over 12 years of age without a documented infection in the registry. The number of uninfected people on June 1^st^ 2022 (the start of the BA.5 dominance period) was 5 328 287, representing 57% of the Portuguese population over 12 (data from the National Census 2021^18^).

The data available in the national COVID-19 registry (*SINAVE*) only include cases of positive tests (PCR tests and rapid antigen tests) performed by healthcare workers in accredited diagnostic facilities. Testing by an accredited facility is a requisite for access to social security compensation for days of isolation – this is a reason for the comprehensiveness of the registry and the exclusive inclusion of validated tests. Only tests performing above the EU-defined minimum for test sensitivity and specificity are used in Portugal. Furthermore, until recently, Portugal had a wide and mandatory testing policy, requiring the presentation of tests for access to several locations, even for vaccinated people (namely, access to entertainment, sports, or healthcare venues).

It is anticipated that the population we classified as “uninfected” included individuals with a prior unnoticed infection. In a previous publication we have shown that the calculated risk of BA.5 infection does not significantly change even if we assume a percentage of 29.2% unreported infections in each period^1^. The number of unreported cases was estimated by the National Serological Panel (from November 2021, assaying the presence of antibodies against the SARS-CoV-2 N protein to exclude vaccination seropositivity) to infer that, at that time, there were 29.2% more people who had been infected with SARS-CoV-2 than officially reported^19^. We further showed that the impact of 20% or 40% of unreported infections within the “uninfected” group did not lead to major changes in the calculated risks^1^.

We used the national SARS-CoV-2 genetic surveillance database^7^ to identify periods when one variant represented >90% of the sample isolates, as also defined and used in other studies ^1,3^. We assigned infected individuals to the variants’ dominance periods and excluded all individuals who had more than one infection before the study period (Figure 1). We pooled BA.1 and BA.2 infections, given the slow transition between the period of dominance of these two subvariants.

Reinfection was defined as two positive tests in the same individual, at least 90 days apart^8^. Consequently, all cases of infection in the 90 days before the start of each study period were not included, as these would not classify as “in risk of reinfection” for the entire duration of the study period under the definition above.

In conclusion, the study design is similar to a prospective study but taking place in the past: the groups of interest were selected (i.e., individuals with one infection in a defined period of time and without any additional infection reported until the start of the study period); and afterwards the individuals from the different groups were followed, under the same epidemiological conditions, for a pre-defined (and equal) number of days and their infections were recorded.

### Vaccination coverage

The vaccine coverage with the primary vaccination series in the Portuguese residents over 12 years was >98% by the end of 2021 ^20^. The primary series of the vaccination campaign used EU/EMA-authorized vaccines: Comirnaty (Pfizer/BioNTech), 69%; Spikevax (Moderna), 12%; Vaxzevria (AstraZeneca), 13%; and Janssen 6%.

While at the start of the BA.1/BA.2 period of dominance (January 1^st^ 2022), the coverage with the first booster was residual (mostly long-term care facility residents), at the start of the BA.5 period of dominance (June 1^st^), the coverage with the first booster was 82%. The vaccine boosters relied exclusively on mRNA vaccines (77% Comirnaty and 23% Spikevax). At the start of the BA.5 period, a second booster was not yet in use except for a highly specific (and small) population of patients with severe immunosuppression.

### Statistics

We analyzed relative risk using the modified Poisson regression method with a robust sandwich estimator for the variance as described^21^. We compared the risk of BA.5 infection for people with a previous single infection at different intervals, with this risk for people without any previous recorded infection. Protection efficacy was estimated, in percentage, as (1-RR) x 100%. Confidence intervals for the RR were calculated using the Wald normal approximation method, with the *epitools* R package^22^. A similar approach was used to calculate the risk of BA.1/BA.2 infection among people previously infected with the Delta variant of SARS-CoV-2.

## Data Availability

Data from the Portuguese COVID-19 registry (SINAVE) is available upon reasonable request from Direcao Geral de Saude Portugal which curates the registry.

## Ethics statement

The study design was approved by the Ethics Review Board of the Centro Academico de Medicina de Lisboa.

